# Ovarian follicular function is not altered by SARS-Cov-2 infection or BNT162b2 mRNA Covid-19 vaccination

**DOI:** 10.1101/2021.04.09.21255195

**Authors:** Yaakov Bentov, Ofer Beharier, Arbel Moav-Zafrir, Maor Kabessa, Miri Godin, Caryn S. Greenfield, Mali Ketzinel-Gilad, Efrat Esh Broder, Hananel E. G. Holzer, Dana Wolf, Esther Oiknine-Djian, Iyad Barghouti, Debra Goldman-Wohl, Simcha Yagel, Asnat Walfisch, Anat Hersko Klement

**Author notes:** Equal Contribution (Yaakov Bentov, Ofer Beharier).

## Abstract

**Importance:** This is the first study to examine the impact of SARS-Cov-2 infection and COVID-19 vaccination on ovarian function.

**Objective:** To characterize anti-COVID-19 antibodies in follicular fluid and compare ovarian follicle function in women following confirmed SARS-CoV-2 infection, COVID-19 vaccination, and non-infected, unvaccinated controls.

**Design:** This is a cohort study conducted between February 1 and March 10, 2021.

**Setting:** A single university hospital-based IVF clinic.

**Participants:** Consecutive sample of female patients undergoing oocyte retrieval.

**Interventions:** Consenting patients were recruited and assigned to one of three study groups: recovering from confirmed COVID 19 (n=9); vaccinated (n=9); and uninfected, non-vaccinated controls (n=14). Serum and follicular fluid samples were taken and analyzed for anti-COVID IgG as well as estrogen, progesterone and HSPG2 concentration, as well as the number and maturity of aspirated oocytes and previous estrogen and progesterone measurements.

**Main outcome measures:** Follicular function, including steroidogenesis, follicular response to the LH/hCG trigger, and oocyte quality biomarkers.

**Results:** Both natural and vaccine elicited anti-COVID IgG antibodies were detected in the follicular fluid in levels proportional to the IgG serum concentration. No differences were detected in any of the surrogate ovarian follicle quality reporting parameters.

**Conclusions and relevance:** Both SARS-COV-2 infection and vaccination with the BNT162b2 mRNA vaccine mediate IgG immunity that crosses into the follicular fluid. No detrimental effect on follicular function was detected.

**Trial Registration:** CinicalTrials.gov registry number NCT04822012

**Key Point:** COVID 19 disease and BNT162b2 mRNA vaccine induce anti-COVID IgG in follicular fluid; neither recent infection nor vaccination appear to negatively effect follicular function.

## Introduction

Since emerging in the last months of 2019, over 127,000,000 individuals have contracted confirmed SARS-CoV-2 infection, and the documented death toll from COVID-19 had reached 2,788,639 individuals globally by the end of March, 2021 ^1^. Jerusalem, the setting of the present study, has presented one of the highest incidence rates of COVID-19 in Israel^2^

Nationwide anti-COVID-19 vaccination began in Israel in December 2020, using the Pfizer – BioNtech vaccine (BNT162b2 mRNA). By the end of March 2021, 5,216,801 and 4,700,957 individuals received the first and second doses of the vaccine, representing 56.1% and 50.55% of Israel’s population, respectively. Vaccination for reproductive age individuals began in January 2021; by the end of March 2021, 72.3% and 61.3% of 20-29 year olds, 77.1% and 67.8% of 30-39 year olds, and 82% and 74.5% of 40-49 year olds had received the first and second doses of the vaccine, respectively, the highest proportions in the world^2^.

Both the rapidly spreading disease and the vaccination campaign were associated with concerns regarding potential detrimental effects on future fertility^3-5^. There is still a lack of real-world data to assist clinicians in counseling their IVF patients regarding the possible impact of recent recovery from COVID infection or vaccination against it, on the potential for success of assisted reproductive treatments. While it has been suggested that COVID-19 might impact fertility, no studies, to the best of our knowledge, have proven a direct gonadal effect of either the disease or the vaccine ^4-8^.

We aimed to determine the impact of confirmed COVID-19 disease and/or immunization on human follicular function, by comparing follicular steroidogenesis, response to the LH/hCG trigger, and oocyte quality biomarker (HSPG2), in the aspirated follicular fluid of patients undergoing ovum pick-up.

## Methods

The study was approved by the Hadassah Medical Center IRB (Permit 0053-21-HMO). The study was registered at the clinical trial registry and assigned the registration number NCT04822012 (Protocol # 0053-21-HMO). Consecutive patients undergoing OPU for either IVF, ICSI, or oocyte cryopreservation were approached to participate in the study on the day of ovum pickup. Eligibility criteria were age older than 18 years and willingness to participate and provide informed consent. Patients younger than 18 or older than 44 years, as well as patients with poor ovarian response (less than 3 mature follicles) were excluded.

After providing informed consent, the patients were questioned about their confirmed past SARS-Cov-2 infection / vaccination status. For recovering patients, the date of the recovery (negative nasopharyngeal COVID PCR test) was recorded. Vaccinated patients were asked about the date of the first and second vaccines. None of the recovering patients were vaccinated with anti-COVID vaccine. The clinic’s strict policy is to screen all patients using a nasopharyngeal swab for COVID-19 a week prior to the procedure, except those patients who were less than 3 months following recovery from SARS-Cov-2 infection, and those more than 2 weeks after the second COVID-19 vaccine. As a result, together with the negative anti-COVID serum antibody test, we could safely presume that all control patients in this study were indeed COVID-19 negative. During the oocyte retrieval the first follicle/s were aspirated into an empty tube. If the sample was contaminated with blood another clean sample was taken, until a total volume of 5 ml of a clean sample was achieved. A 5 ml blood sample was also taken during the procedure. Following the isolation of the oocyte, the FF was centrifuged at 1500g and the blood sample at 3000g for 7 minutes. The supernatant fraction of the follicular fluid and the serum fraction of the blood sample were each aliquoted and later snap frozen and stored at −80 C° until analysis.

Data including the patient age, IVF indication, antral follicle count (AFC), serum estradiol and progesterone on the day of ovulation trigger (36 hours before oocyte retrieval), type of trigger, the number of oocytes, and mature oocytes, were recorded.

Once the target date was reached the samples were thawed and analyzed for the following assays. The analysis of the blood and FF samples for all outcome parameters was conducted with blinding of the COVID / Vaccine status of the participant.

### Serum and follicular fluid anti-COVID IgG measurement

The levels of specific anti-SARS–CoV-2 spike protein receptor binding domain (RBD) IgG were assessed in serum and follicular fluid specimens, using the Architect SARS-CoV-2 IgG II Quant assay (Abbott Diagnostics, Chicago, USA), according to the manufacturer’s specifications. IgG levels ≥ 50 AU/mL were considered positive.

### Assessment of ovarian follicle functions

#### Steroidogenesis

We examined the ability of the Theca-granulosa cells that form the wall of the follicles to produce steroids, namely estradiol and progesterone, by measuring their concentration both in the serum and FF.

The measurement was conducted using the Atellica IM Siemens Healthineers system (Siemens Healthcare GmbH Henkestr. 127 91052 Erlangen Germany). Estradiol concentration was measured using the Enhanced Estradiol Kit (# 10995561) an ELISA based on acridinium-labeled sheep monoclonal anti-estradiol antibody with a measuring range of 43.31-11,010.0 pmol/L). For measuring FF estradiol, the typical concentration of which exceeds the measuring range of the kit, the sample was diluted using the Atellica IM eE2 diluent (10995563) according to a protocol described elsewhere ^9^. The measurement of serum and FF progesterone was conducted using the PRGE kit (10995660). The kit is based on direct chemiluminescent technology using an acridinium-labeled mouse monoclonal anti-progesterone antibody with a measuring interval of 0.67-190.80 nmol/L. For FF progesterone measurement, the sample was diluted with the Atellica IM Multi-diluent (10995645) according to the protocol ^9^.

To normalize estradiol production, we used the ratio of serum estradiol on the day of the trigger per retrieved oocyte. This ratio was found in a large study^10^ to be an age independent predictor of IVF treatment outcome. In that study an estradiol/oocyte ratio of > 1,000 pmol/L/Oocyte was associated with a reduction in pregnancy and clinical pregnancy rate^10^.

#### Follicular response to the LH/hCG trigger assessment

To assess the adequacy of response of the follicle to the LH/hCG trigger we compared the following parameters:

1. Oocyte yield: the ratio between the number of retrieved oocytes to the number of mature follicles measured on the day of the trigger injection. This ratio represents the ability of the granulosa cells to secrete enzymes that release the cumulus oocyte complex from the wall of the follicle in response to the LH surge, thereby allowing its aspiration. An oocyte yield of 45% or less was defined as sub-optimal ^11^.

2. Ratio of mature oocytes to total aspirated oocytes, representing another aspect of the adequacy of response to the LH/hCG trigger – promoting the completion of the first meiotic division.

3. Ratio of the number of retrieved oocytes to the number of follicles that began luteinisation, obtained by dividing the number of retrieved oocytes by the serum concentration of progesterone on the day of OPU.

#### Assessment of oocyte quality FF biomarkers

Concentration of Heparan Sulphate Proteoglycan II in follicular fluid (HSPG2): A proteomic analysis of over 500 FF proteins identified the concentration of HSPG2 (Pearlecan) in FF as the strongest biomarker to predict oocyte fertilization and IVF success ^12^.

We analyzed the concentration of HSPG2 in FF using the Human HSPG2 SimpleStep ELISA kit (Abcam, Discovery Drive, Cambridge Biomedical Campus, Cambridge, CB2 0AX, UK). This ELISA system uses an affinity tag labeled capture antibody and a reporter conjugated detector antibody read at 450-600nm. The test range of measurement is 47 −7,000 pg/ml. Dilution of the samples with up to 1:50k was done with a proper diluent. The analysis was conducted according to the manufacturer’s instructions.

### Statistical analysis

All analyses were performed using SPSS 23.0 (SPSS Inc., Chicago, IL). Continuous intervals and ratios that were not normally distributed were transformed to a logarithmic scale for normality correction. Normally distributed data were compared across study groups by univariate ANOVA. Rates and proportions were compared with the Chi-square or Fisher’s exact tests as appropriate in case of small numbers. Correlations were presented by scatter plots and the goodness of fit calculated by R^2^. All P values were tested as two tailed and considered significant at <0.05.

## Results

### Patient characteristics

Overall, 32 patients consented to participate in the study, of which 9 reported vaccination (Group 1 – Vaccine); 7 to have recovered from COVID (Group 2 – disease); and 16 denied both and were allocated to the non-exposed group (Group 3). However, serum analysis for the presence of anti-COVID-19 IgG identified two of the patients in the control group as recoverees from a likely asymptomatic SARS-Cov-2 infection and therefore were re-allocated to Group 2. Mean time periods from vaccine or recovery to recruitment and sampling were as follows: 93.2 days from recovery to sampling (range 48-169 days); 11.7 days from first vaccine dose to sampling in the 4 patients who were recruited after completion of a single vaccine dose (range 8-18 days) and 27.6 days from second vaccine dose to sampling in 5 patients recruited after completing two doses (range 4-46 days). Patient demographics are described in Table 1.

**Table 1:** Patient’s demographics by study group.

|  |  | Vaccine | Disease | Non exposed | Total |  |
| --- | --- | --- | --- | --- | --- | --- |
| Number |  | 9 | 9 | 14 | 32 | P Value |
| Age | Mean ± *STDEV | 35.3 ± 3.97 | 34.1 ± 4.7 | 32.5 ± 5.3 | 33.75 (± 4.8) | 0.383 |
|  | Median | 35 | 34 | 33 | 34 |  |
| Antral follicular count | Mean ± STDEV | 13.3 ± 4.7 | 13.6 ± 4.1 | 15.6 ± 6.7 | 14.4 ± 5.7 | 0.592 |
|  | Median | 14 | 13 | 17.5 | 14.5 |  |
| Indication | Male (%) | 2 (22.2%) | 1 (11.1%) | 4 (28.6%) | 7 (21.9%) | 0.113 |
|  | Non male (%) | 2 (22.2%) | 5 (55.6%) | 9 (64.3%) | 16 (50%) |  |
|  | Egg Freezing (%) | 5 (55.6%) | 3 (33.3%) | 1 (7.1%) | 9 (28.1%) |  |
|  | Total non-Infertile | 7 (77.8%) | 4 (44.4%) | 5 (35.7%) | 16 (50%) |  |
| Time interval(days) | Mean | 32.2 | 98.14 | N/A |  | 0.003 |
|  | STD DEV | 22.1 | 45.5 | N/A |  |  |

\*STDEV: standard deviation

The mean age of participants in the study was 33.7 years. Age was found to be normally distributed and did not vary significantly among the 3 groups (_P_ value 0.383). Similarly, measurement of antral follicle count, as a reflection of ovarian reserve, did not differ among the groups (*P* value 0.592). Indications for treatment were grouped into non-male related infertility, male related infertility, and oocyte cryopreservation. The latter two groups therefore included women with no proven infertility. Only women in the first two groups had their oocytes fertilized following the oocyte pick up (OPU). There were more women undergoing OPU for oocyte freezing in the vaccine group (55.6%) compared to the other groups (33.3% and 7.1%): this difference was close to statistical significance (*P* value 0.07). In all groups a large proportion of women had OPU for non-female infertility related indications (50%, range 35.7-77.8%). As most patients participating in the study were treated with an antagonist protocol, the use of hCG only trigger was limited (6.3%). The use of GnRH agonist trigger reflected the distribution of OPU for oocyte cryopreservation in the different groups (44%, 44%, and 28.6% in the vaccinated, COVID, and control groups, respectively, P value 0.766).

### Anti-COVID IgG antibodies

Table 2 describes the presence of anti-COVID IgG induced by either SARS-COV-2 infection or secondary to vaccination with the Pfizer – BioNtech vaccine (BNT162b2 mRNA). All the patients who received two doses of the BNT162b2 mRNA vaccine had a high concentration of COVID IgG in their serum and FF (range 850-19,672 and 463-15,883 in the serum and FF respectively). IgG concentrations were positively correlated with the time interval from the date of the vaccine: two of the patients that had only received the first dose, 13 and 18 days before OPU, were found to harbour anti-COVID IgG in their serum and FF (range 194-228 and 64-198 in the serum and FF respectively), while the two who received the first dose only 8 days prior to OPU had no measurable anti-COVID IgG either in their serum or FF. All the patients with positive serum anti-COVID IgG had detectable levels of anti-COVID IgG in their follicular fluid. Figure one shows a linear association between the serum and FF concentrations of anti-COVID IgG (R^2^ =0.99), suggesting a nonregulated passage of the anti-COVID IgG across the follicular membrane.

**Table 2:**
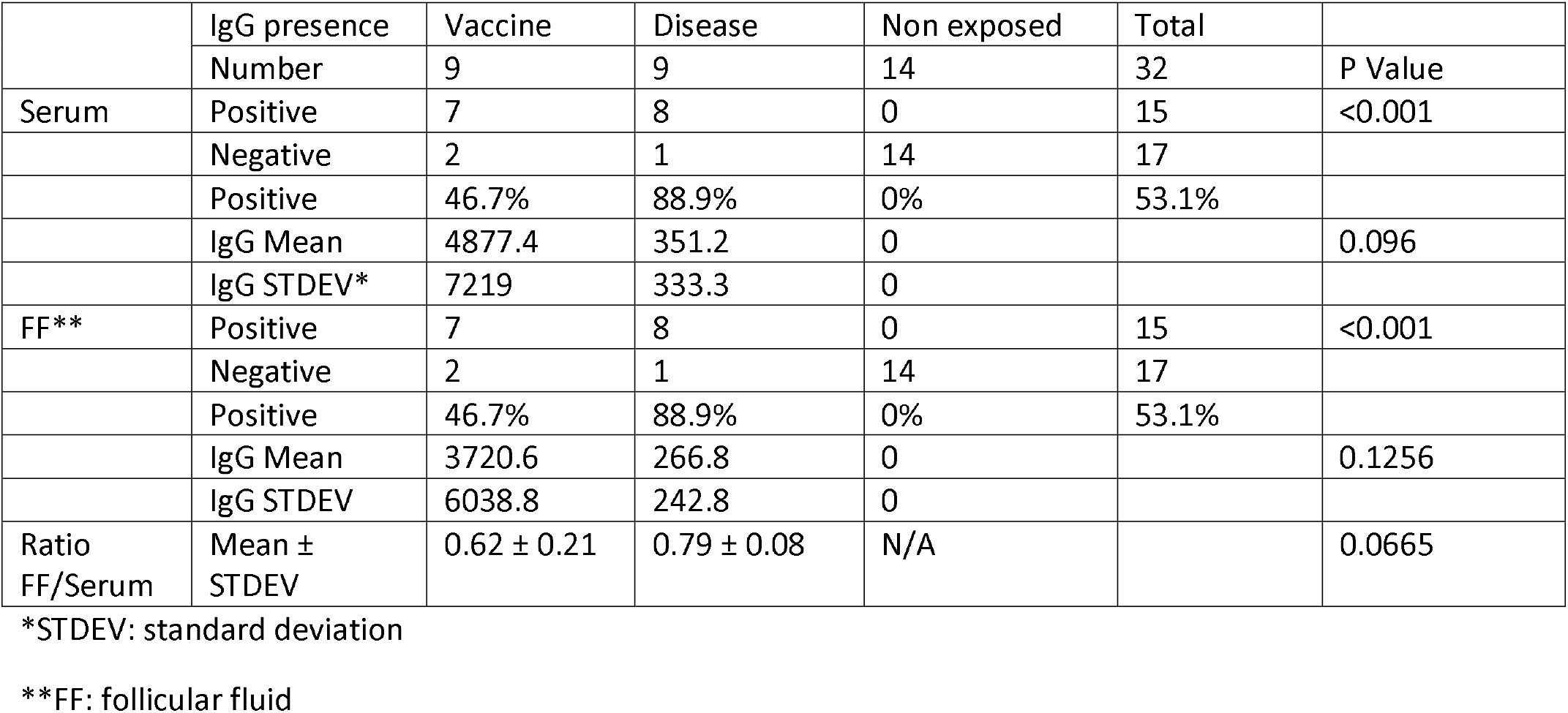
Serum and Follicular Fluid IgG level by study group.

No difference was found between women acquiring anti-COVID antibodies following exposure or vaccination, in the ratio between FF to serum anti-COVID IgG.

### Assessment of ovarian follicle functions

Table 3 describes treatment and outcome parameters. These parameters provide data on the steroidogenic function of the follicle (trigger day serum and FF estradiol and progesterone concentrations) and the magnitude of the ova recruitment, as reflected by the number of aspirated eggs.

**Table 3:**
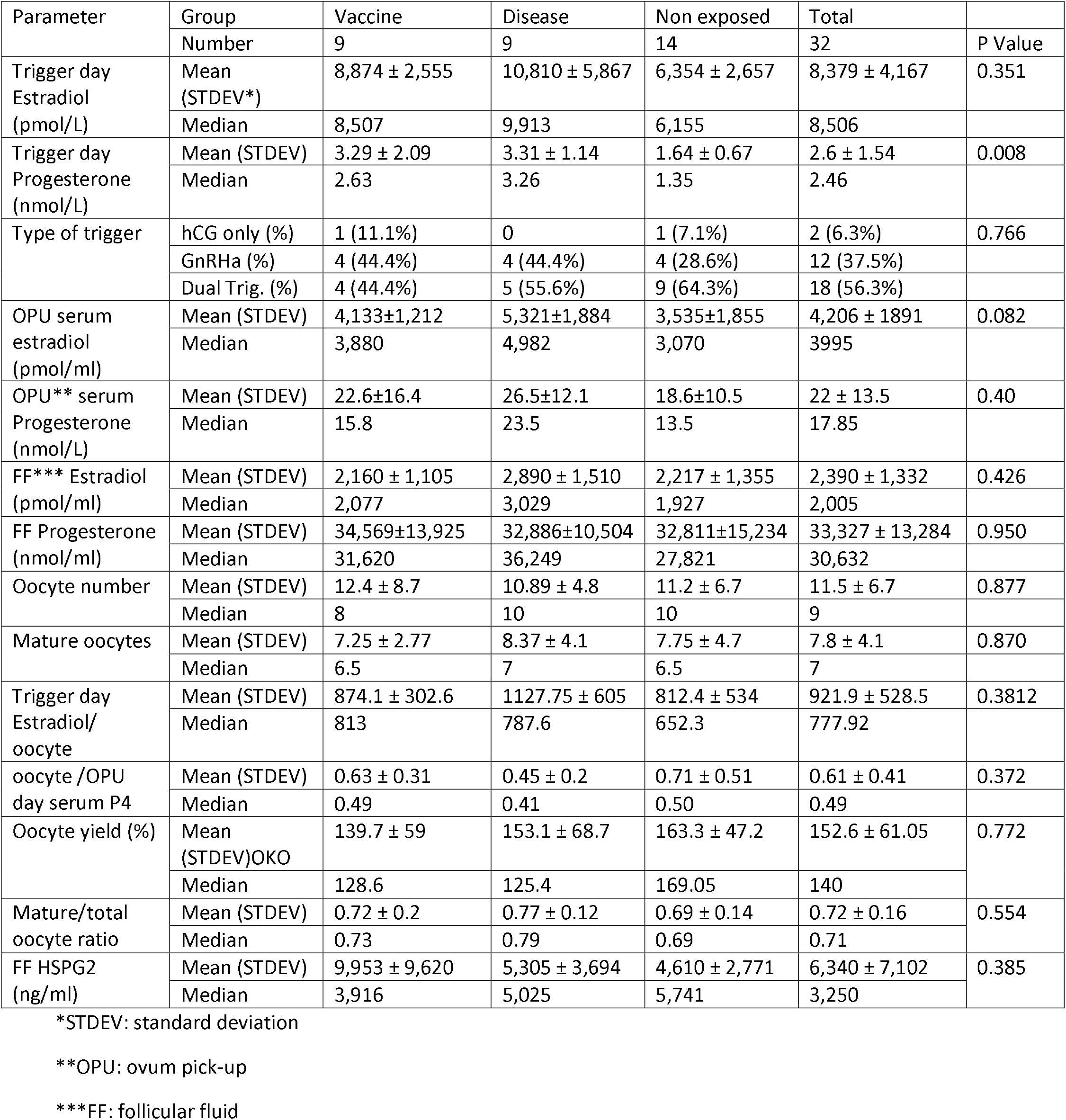
Treatment parameters by study group.

| Parameter | Group | Vaccine | Disease | Non exposed | Total |  |
| --- | --- | --- | --- | --- | --- | --- |
|  | Number | 9 | 9 | 14 | 32 | P Value |
| Trigger day Estradiol (pmol/L) | Mean (STDEV*) | 8,874 ± 2,555 | 10,810 ± 5,867 | 6,354 ± 2,657 | 8,379 ± 4,167 | 0.351 |
|  | Median | 8,507 | 9,913 | 6,155 | 8,506 |  |
| Trigger day Progesterone (nmol/L) | Mean (STDEV) | 3.29 ± 2.09 | 3.31 ± 1.14 | 1.64 ± 0.67 | 2.6 ± 1.54 | 0.008 |
|  | Median | 2.63 | 3.26 | 1.35 | 2.46 |  |
| Type of trigger | hCG only (%) | 1 (11.1%) | 0 | 1 (7.1%) | 2 (6.3%) | 0.766 |
|  | GnRHa (%) | 4 (44.4%) | 4 (44.4%) | 4 (28.6%) | 12 (37.5%) |  |
|  | Dual Trig. (%) | 4 (44.4%) | 5 (55.6%) | 9 (64.3%) | 18 (56.3%) |  |
| OPU serum estradiol (pmol/ml) | Mean (STDEV) | 4,133±1,212 | 5,321±1,884 | 3,535±1,855 | 4,206 ± 1891 | 0.082 |
|  | Median | 3,880 | 4,982 | 3,070 | 3995 |  |
| OPU** serum Progesterone (nmol/L) | Mean (STDEV) | 22.6±16.4 | 26.5±12.1 | 18.6±10.5 | 22 ± 13.5 | 0.40 |
|  | Median | 15.8 | 23.5 | 13.5 | 17.85 |  |
| FF*** Estradiol (pmol/ml) | Mean (STDEV) | 2,160 ± 1,105 | 2,890 ± 1,510 | 2,217 ± 1,355 | 2,390 ± 1,332 | 0.426 |
|  | Median | 2,077 | 3,029 | 1,927 | 2,005 |  |
| FF Progesterone (nmol/ml) | Mean (STDEV) | 34,569±13,925 | 32,886±10,504 | 32,811±15,234 | 33,327 ± 13,284 | 0.950 |
|  | Median | 31,620 | 36,249 | 27,821 | 30,632 |  |
| Oocyte number | Mean (STDEV) | 12.4 ± 8.7 | 10.89 ± 4.8 | 11.2 ± 6.7 | 11.5 ± 6.7 | 0.877 |
|  | Median | 8 | 10 | 10 | 9 |  |
| Mature oocytes | Mean (STDEV) | 7.25 ± 2.77 | 8.37 ± 4.1 | 7.75 ± 4.7 | 7.8 ± 4.1 | 0.870 |
|  | Median | 6.5 | 7 | 6.5 | 7 |  |
| Trigger day Estradiol/oocyte | Mean (STDEV) | 874.1 ± 302.6 | 1127.75 ± 605 | 812.4 ± 534 | 921.9 ± 528.5 | 0.3812 |
|  | Median | 813 | 787.6 | 652.3 | 777.92 |  |
| oocyte /OPU day serum P4 | Mean (STDEV) | 0.63 ± 0.31 | 0.45 ± 0.2 | 0.71 ± 0.51 | 0.61 ± 0.41 | 0.372 |
|  | Median | 0.49 | 0.41 | 0.50 | 0.49 |  |
| Oocyte yield (%) | Mean (STDEV)OKO | 139.7 ± 59 | 153.1 ± 68.7 | 163.3 ± 47.2 | 152.6 ± 61.05 | 0.772 |
|  | Median | 128.6 | 125.4 | 169.05 | 140 |  |
| Mature/total oocyte ratio | Mean (STDEV) | 0.72 ± 0.2 | 0.77 ± 0.12 | 0.69 ± 0.14 | 0.72 ± 0.16 | 0.554 |
|  | Median | 0.73 | 0.79 | 0.69 | 0.71 |  |
| FF HSPG2 (ng/ml) | Mean (STDEV) | 9,953 ± 9,620 | 5,305 ± 3,694 | 4,610 ± 2,771 | 6,340 ± 7,102 | 0.385 |
|  | Median | 3,916 | 5,025 | 5,741 | 3,250 |  |

\*STDEV: standard deviation
\*\*OPU: ovum pick-up
\*\*\*FF: follicular fluid

Mean serum estradiol on the day in which the trigger injection was administered (36 hours prior to OPU) did not differ among the groups (P value 0.351). The normalized ratio of serum estradiol on trigger day / oocyte was similar and within the optimal range for most of the patients in all groups. Serum progesterone on the same day was lower in the non-exposed group (3.29, 3.31 and 1.64 nmol/L in the vaccine, COVID and non-exposed respectively, P value 0.008) however no difference was measured in serum progesterone on the day of OPU (22.6, 26.5 and 18.6 nmol/L in the vaccine, COVID and control groups respectively, P value 0.40).

Although both FF estradiol and progesterone concentrations were hundreds fold higher compared to serum concentrations on the same day, no difference was found among the groups (2,160, 2,890 and 2,217 nmol/L for estradiol and 34,569, 32.886 and 32,811 nmol/L for progesterone in the vaccine, COVID and non-exposed groups, respectively, P values 0.426 and 0.950).

The adequacy of the follicular response to ovulation triggering was assessed by serum and FF progesterone as well as by the oocyte yield (oocytes retrieved/trigger day mature follicle count, oocytes retrieved /OPU day serum progesterone, ratio of mature/ total number of aspirated oocytes).

Oocyte yield was high in all groups (mean 152%, P value 0.772) well above the 45% threshold defining a sub-optimal response. Similarly, the rate of mature oocytes was normal (mean 0.72 P Value 0.554). The ratio between the number of aspirated oocytes and OPU day progesterone was 0.63, 0.45, and 0.61 in the vaccine, COVID, and control groups respectively, P value 0.372.

### Assessment of oocyte quality with follicular fluid HSPG2

None of these follicle quality reporter surrogates suggested any meaningful differences among the 3 groups. The mean FF HSPG2 for all samples was 6,340 ng/ml (median 3,250, range 302-43,603). Comparison of HSPG2 between the groups showed similar concentrations (*P* Value 0.385).

## Discussion

Recent publications have indicated a potential for severe morbidity and mortality among parturients affected by the emerging variants of the SARS-COV-2 virus ^13^. Despite the low incidence of severe morbidity among parturients affected by COVID 19, the CDC added pregnancy to the list of high-risk conditions to prioritize vaccination, and ACOG recommends not withholding vaccination from pregnant women at any stage of the pregnancy ^14^. The enthusiasm surrounding the vaccine rollout was accompanied by unsubstantiated rumors spread via social media, suggesting that the vaccine may lead to female sterility^4^. The risk for severe COVID related illness on one hand and the lack of knowledge about the potential effects of anti-COVID vaccination, led to much apprehension among patients planning to conceive, resulting in couples postponing their plans to conceive^3 13^. To the best of our knowledge, our study is the first to determine the ovarian involvement with the COVID immune response, as reflected by the presence of anti-SARS-COV-2 IgG in recently vaccinated vs. infected and non-vaccinated non-infected IVF patients, all of whom were PCR negative at the time of OPU.

Our results show that all the patients reported as COVID-19 recoverees had measurable anti-SARS-COV-2 IgG both in their serum and FF. The presence of anti-COVID IgG following vaccination was shown as early as 13 days following the first dose, both in the serum and FF. The concentration of anti-COVID IgG correlated with the time interval from the vaccine. The anti-COVID IgG in the FF reflects its serum concentration in a linear association, as previously described for other vertically transmitted viruses ^15^.

Our assessment of a potential effect of either infection with SARS-COV-2 or vaccination on ovarian function included several aspects related to the egg-follicle performance. Assessment of follicular steroidogenesis showed similar and normal rates of estrogen and progesterone production. The only significant difference we detected was a lower trigger day serum progesterone in the non-exposed group, a difference that was not shown on the day of OPU, either in the serum or FF, and therefore is unlikely to represent a real biological difference. Otherwise, none of the synthetic parameters studied showed any differences among the 3 groups. Assessment of the response of the follicles to the LH/hCG trigger showed a normal and similar response in all 3 groups.

### Oocyte quality assessment

We compared the concentration of FF HSPG2, a validated biomarker of oocyte quality. FF samples showed a high concentration of HSPG2 that was not significantly different between the groups.

Hence, despite the clear evidence of intimate follicular immune exposure post infection with SARS-COV-2 or following BNT162b2 mRNA vaccine, the steroidogenic machinery of the follicle, controlling the ultimate maturation of the oocyte and its hormonal milieu, did not show any measurable difference as compared to non-exposed women. This evidence joins other data on IVF treatment outcomes among recoverees from COVID-19 infection observed in our center, which showed similar outcomes (data not shown).

### Strengths and limitations

To the best of our knowledge this is the first study to examine the presence of natural or vaccine-elicited humoral immune response in the ovarian follicle. In addition, we also investigated the possible association between its presence and several aspects of follicular function and oocyte yield. We chose not to examine the rate of fertilization and embryo formation and development, as they involve the male gametes and may introduce differences unrelated to the study question.

This is however a small study, comprising a mixed fertile and infertile population, and its conclusions should be supported and validated by larger studies. Due to its timing, our study was able to assess short term affects of vaccine and disease but cannot rule out later sequelae.

## Conclusions

Exposure to either the SARS-COV-2 virus or the BNT162b2 mRNA vaccine results in a rapid formation of anti-COVID IgG that can be detected in the follicular fluid, correlating with serum concentrations. In our study, neither infection or the BNT162b2 mRNA vaccine, nor the immune response to them, resulted in any measurable detrimental effect on the function of the ovarian follicle.

## Data Availability

All data is available upon a reasonable request

---

**Figure 1:**
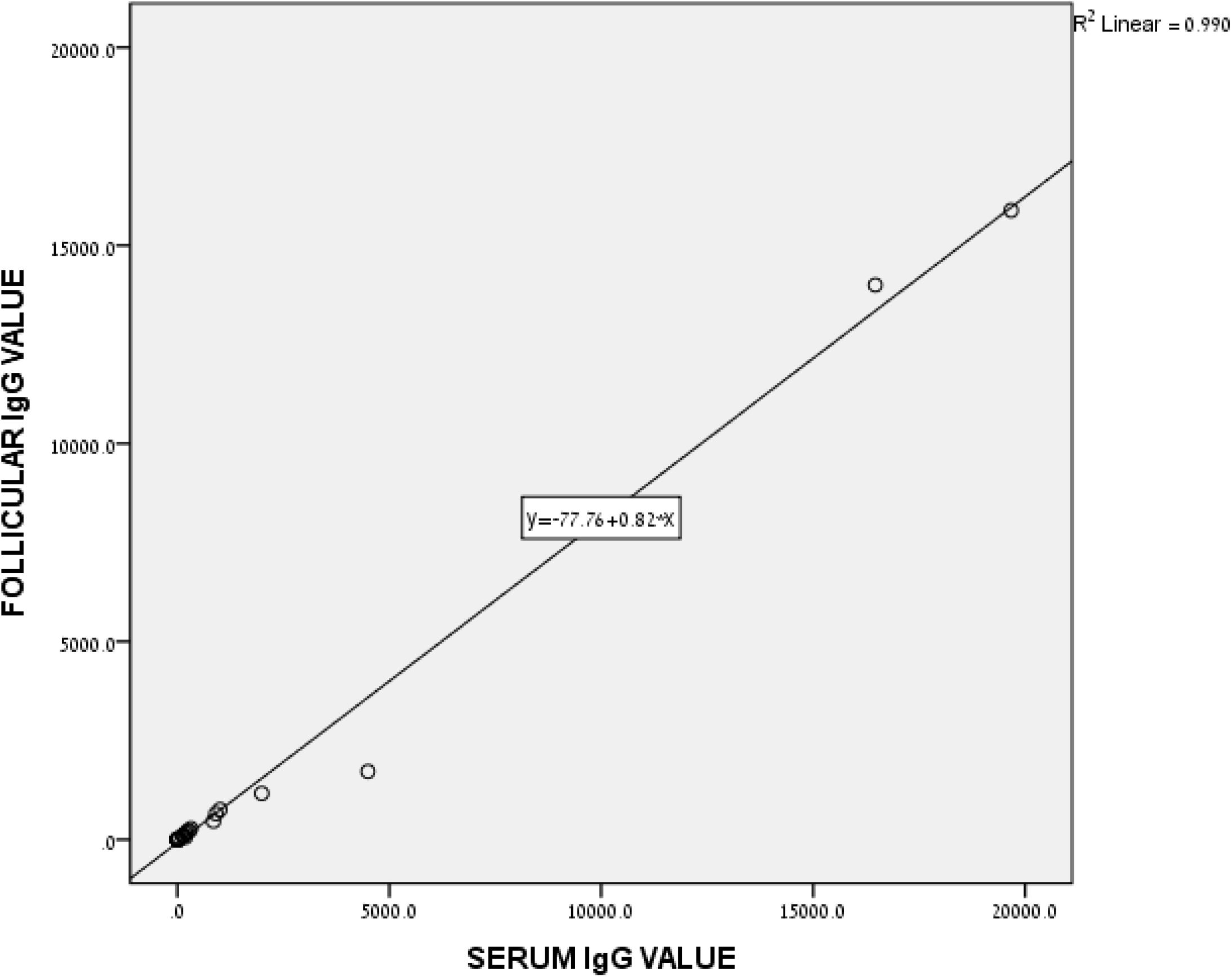
The relationship of follicular fluid and serum anti-SARS-COV-2 IgG concentration

